# Latent biomarker states underlying disagreement between PET-anchored and distribution-based plasma pTau-217 positivity thresholds

**DOI:** 10.64898/2026.07.17.26358314

**Authors:** Kalliopi Mavromati, Adam H. Dyer, Jack D. Beazer, Lynne Hughes, Sean P Kennelly, Terence J Quinn

**Author notes:** **Corresponding author:** Kalliopi Mavromati.

## Abstract

**Background:** Plasma phosphorylated tau-217 (pTau-217) measurements for use in Alzheimer’s disease (AD) identification require thresholds to define positivity and there exist different approaches to operationally defining the boundary. We compared amyloid β (AB) PET-anchored and distribution-based positivity cut-off values and explored how these mapped onto latent biomarker states.

**Methods:** We analysed plasma pTau-217 measured in the Bio-Hermes-001 cohort (N = 990) using an immunoassay (Lilly) and mass spectrometry assay (University of Gothenburg). Gaussian mixture models were used to identify latent classes and thresholds were derived in two ways: achieving 90% specificity for AB PET positivity and exceeding the mean + 2SDs of the lowest latent class. We explore classes in reference to AB PET status and clinical diagnosis, as well as agreement between approaches using Cohen’s kappa for both assays.

**Results:** In both assays, three latent biomarker classes were identified with monotonic increases in AD clinical diagnosis and AB PET positivity. PET-anchored thresholds showed lower specificity but higher sensitivity to amyloid positivity than distribution-based thresholds. Overall agreement between the approaches was acceptable (k = 0.678 for Lilly and 0.575 for University of Gothenburg), with disagreement concentrated in the intermediate latent class. Classes with the lowest and highest pTau-217 concentrations were classified consistently using both thresholds

**Discussion:** The two thresholding approaches yielded similar classifications at both the negative and positive tail of the observed biomarker distribution, but classify intermediate concentrations differently. The boundary definition influenced pTau-217 positivity more than the analytical platform itself. Thresholding approaches may capture different pTau-217 biomarker states, therefore such methodological decisions should be grounded in the context of the intended application.

## Introduction

Blood phosphorylated tau-217 (pTau-217) has emerged as one of the most promising blood-based biological markers for Alzheimer’s Disease (AD)^1,2^. With strong associations with both cerebral amyloid and tau pathology^2,3^, pTau-217 performs similar to imaging and cerebrospinal fluid (CSF) measures^1,4^. However, unlike categorical clinical and image-derived classifications, fluid biomarkers require thresholds^1,5^ for positivity and negativity built on externally derived reference standards, a process which is still currently ongoing for pTau-217^1,4^. This can be attributed to differences related to detection methodology (e.g. mass spectrometry or immunity-based assays) and proprietary artefacts of the assay (e.g. type of antibody used in immunoassays)^1,3^.

Providing an operational definition for pTau-217 positivity is integral for use of the biomarker in clinical research and practice. It is thus imperative to understand how thresholds relate to markers of clinical and pathologic state. One approach relies on anchoring the blood-based measure to amyloid β (AB) PET^5,6^, the current in vivo gold standard for cerebral amyloid pathology^7^. Another approach is to ground the threshold to the observed blood-based biomarker distribution, without input from clinical presentation or other biomarker modalities. However, in the case of pTau-217, the two approaches may not necessarily converge in identifying positivity. The AB PET-anchored approach operates as a blood-based proxy for AB accumulation in the brain, while the distribution-based threshold inherently reflects the biomarker concentrations of the populations from which it is derived. Moreover, pathogenic staging varies depending on methodology used (e.g. Braak and Braak staging^8^), raising the question of whether alternative operational definitions of pTau-217 positivity converge in reflecting the same state on the biological continuum, despite potentially distinct numerical thresholds.

The operational approach to thresholding a continuous biomarker can have far-reaching implications for any process relying on the classification itself. That includes estimating prevalence of a disease, recruiting participants for both observational and interventional clinical trials, as well as interpreting associations between biomarkers and clinical outcomes^1,7,9^. Given that pTau-217 associations with other biomarkers and clinical outcomes are still being mapped across cohorts and populations^4^, it remains uncertain how different approaches to operationalising pTau-217 positivity could shape what pathologic states the biomarker best reflects.

Using an immunoassay and a mass spectrometry assay from the cross-sectional Bio-Hermes-001, we examined latent biomarker structure using Gaussian mixture modelling and evaluated agreement between PET-anchored and distribution-based thresholds of pTau-217 positivity. In addition to latent biomarker-based states, we explored how the cut-offs mapped onto amyloid PET status, and clinical diagnosis to interpret pathologic states captured by each approach.

## Methods

### Participants

We utilised the cross-sectional Bio-Hermes-001 dataset^10^, hosted on the AD workbench^11^ and accessed via the Data Challenge which also provided ethical approval for using the dataset. Bio-Hermes-001 recruited community-dwelling adults aged 60-85, intentionally engaging participants from typically underrepresented populations (URP) in dementia research. Cognitive and functional screening as well as blood sampling was undertaken during the first of three visits. Where participants could access certified PET facilities, the second visit included an AB PET scan. Then investigators diagnosed participants as healthy cognition (HC), mild cognitive impairment (MCI), or probable mild Alzheimer’s disease (AD) according to National Institute on Aging – Alzheimer’s Association criteria^5^ without incorporating biomarkers^10^.

### Variables

Demographic variables included age and sex. We also utilised the clinical diagnosis (HC/MCI/AD) and the AB PET binary outcome (positive/negative). The original study report that all PET scans from certified centres were reviewed by one team^10^, therefore the methodology used to derive positivity based on PET is consistent across the dataset. N = 54 participants did not have AB PET data due to lack of access to certified PET centres. We did not impute other measures for this missing data. For pTau-217, we utilised the Lilly^12^ and University of Gothenburg (UoG) pTau-217^3^ assays available in the dataset, as accessed through the Data Challenge. The Lilly assay is a Single Molecule Array (Simoa, u/mL), whereas UoG is derived via mass spectrometry (ng/L).

### Statistical Analysis

Analyses were performed on participants with plasma pTau-217 data who completed the study and thus had received a clinical diagnosis (N = 990). The Lilly assay contained entries below the lower limit for detection for which we imputed 0 (N = 174) and above the upper limit for which we imputed the maximum value reported in the cohort (N = 7, max pTau-217 Lilly = 1.39). Ptau-217 concentrations were log-transformed. To explore pTau-217 distributions in the sample, we utilised kernel density plots, grouped by assay as well as clinical classifications. Gaussian mixture models (GMMs)^13^ were fitted to the log-transformed pTau-217 panel results to identify biomarker states according to each assay, independent of clinical diagnosis. We report Bayesian Information Criterion (BIC) for these models. In this manner, participants were assigned to latent classes according to maximum posterior probability derived from the model. We examined the latent classes using uncertainty estimation to quantify the overlap between the latent classes and associations with clinical diagnosis and AB PET informed interpretation of what the classes represented. Chi squares with Cramer’s V effect size were used to quantify associations with clinical diagnosis and AB PET.

Two thresholding approaches to defining thresholds for pTau-217 assays were evaluated. First, the AB PET-anchored approach required that pTau-217 threshold retain at minimum 90% specificity^14^ for distinguishing AB PET positivity. Then, the distribution-based approach^15^ was derived from the lowest GMM-identified latent biomarker class, defining positivity as concentrations exceeding the mean + 2*SD of that class. These methods were implemented to both the Lilly and UoG assays. We report specificity and sensitivity to AB PET classification for pTau-217 thresholds according to both approaches for each assay.

Agreement between the PET-anchored and the distribution-based definitions was quantified using Cohen’s kappa coefficients. Threshold classifications were subsequently examined across latent biomarker classes, AB PET status, and clinical diagnosis to interpret the state along the biological continuum represented by each thresholding approach.

All analyses were conducted using R v4.6.0 ^16^ in RStudio v202.06.0 ^17^ with packages *tidyverse*^18^, *plotROC*^19^, *ggpubr*^20^, *pROC*^21^, *DescTools*^22^, *Resource Selection*^23^, *rms*^24^, *broom*^25^, and *grateful*^26^. All p-values are reported exact, unless <.001.

## Results

This analysis included N = 990 participants with blood biomarker data and complete participation in the study. Individuals were on average 72.1 years of age (SD = 6.66). Over half of the sample were female (N = 559, 56.46%, male: N = 431), and the majority were White (N = 847, 85.55%; Black or African American: N = 113, 11.41%; Asian: N = 19, 1.91% ; American Indian or Alaska Native, or Native Hawaiian or other Pacific Islander, or unknown: N = 11, 1.11%). All clinical diagnosis groups were represented in this sample (HC: N = 416, 42.02%; MCI: N = 308, 31.11%; AD: N = 266, 26.86%).

The assays produced different log-transformed distributions of pTau-217 concentration, with the Lilly SIMOA assay demonstrating a strongly right skew compared to the mass spectrometry UoG equivalent (Figure 1, see also Supplementary Materials). The GMM identified 3-group models as the optimal probabilistic assignment of participants using both assays (Lilly: BIC = -886; UoG: BIC = -1672), with pTau-217 concentration increasing with each class (Table 1).

**Table 1.** Descriptive statistics for each assay by latent class.

| Assay | Latent Class | Count (proportion) | Mean | Mean (SD) uncertainty | Median uncertainty |
| --- | --- | --- | --- | --- | --- |
| Lilly (Simoa, U/mL) | 1 | 327 (35.79) | -1.90 | 0.16 (0.11) | 0.11 |
|  | 2 | 263 (32.08) | -1.56 | 0.24 (0.12) | 0.19 |
|  | 3 | 226 (32.13) | -0.81 | 0.07 (0.13) | 0.002 |
| UoG (Mass Spec, ng/L) | 1 | 486 (48.63) | 0.46 | 0.09 (0.14) | 0.02 |
|  | 2 | 369 (37.11) | 1.16 | 0.17 (0.13) | 0.13 |
|  | 3 | 135 (14.26) | 1.92 | 0.12 (0.13) | 0.06 |

*Note.* Uncertainty refers to posterior uncertainty.

**Figure 1.**
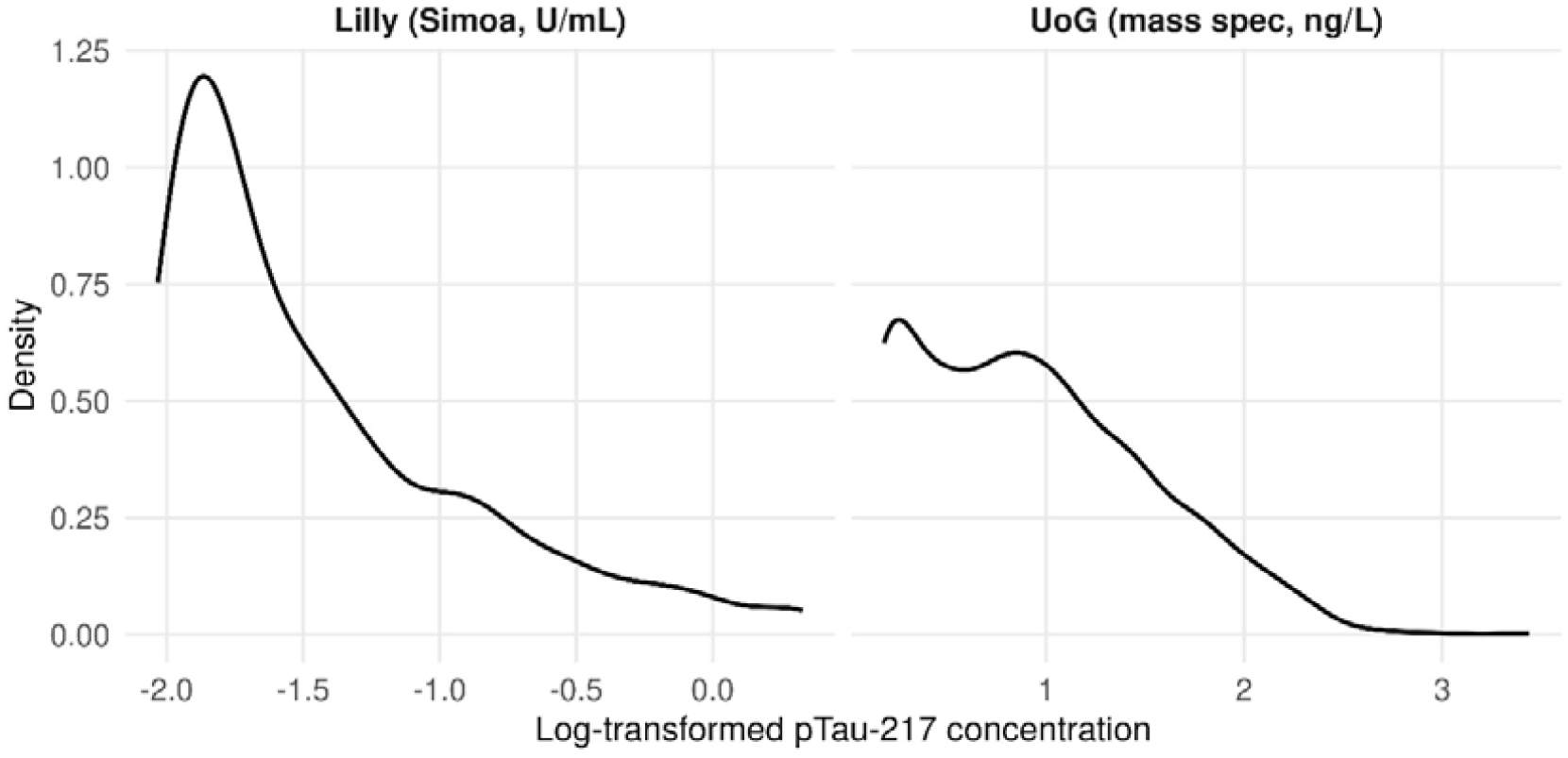
Probability density distribution plots of log transformed pTau-217 concentrations by assay.

The progressive pTau-217 concentration from class 1 to 3 in both assays was moderately positively associated with clinical diagnosis and AB PET positivity (Figure 2, Supplementary Table X, Lilly: with clinical diagnosis χ^2^ (4) = 165, p < .001, Cramer’s V = 0.318 and with AB PET χ^2^ (2) = 331, p < .001, Cramer’s V = 0.654; UoG: with clinical diagnosis χ^2^ (4) = 102, p < .001, Cramer’s V = 0.227 and χ^2^ (2) = 284, p < .001, Cramer’s V = 0.551).

**Figure 2.**
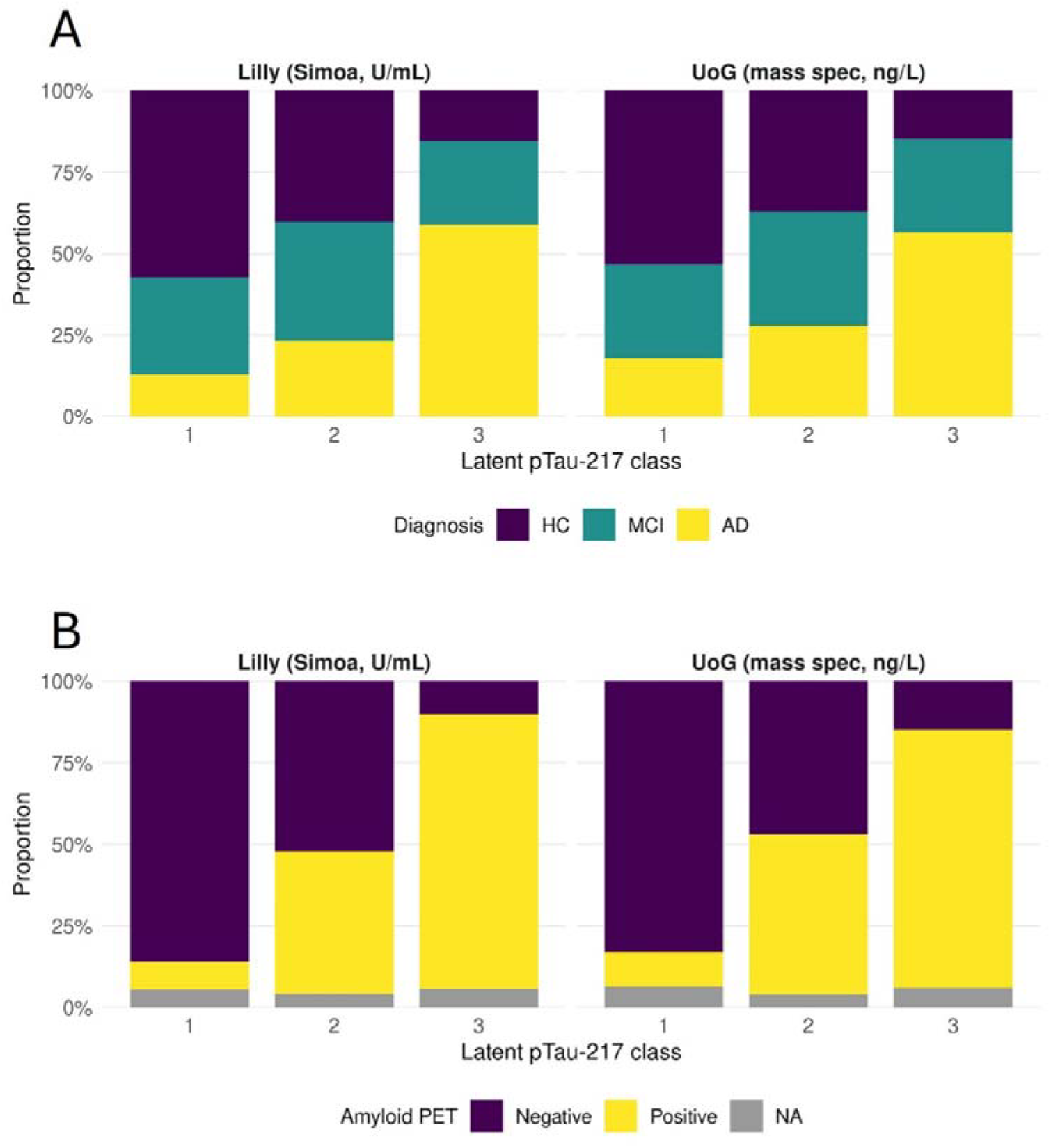
Proportion of latent class by diagnosis and amyloid PET positivity for each assay. *HC* = Healthy Cognition; *MCI* = Mild Cognitive Impairment; *AD* = Alzheimer’s Disease.

According to the PET-anchored approach, we were able to identify pTau-217 thresholds that met the 90% AB PET specificity prespecified requirement (Lilly: threshold = 0.222, specificity = 0.903, sensitivity = 0.799; UoG: threshold = 3.58, specificity = 0.903; sensitivity = 0.584). Calculated according to the distribution-based approach, pTau-217 thresholds greater than the mean + 2*SD had comparatively lower AB PET specificity (Lilly: M = -1.90 (SD = 0.08), threshold = - 1.74, specificity = 0.655, sensitivity = 0.907; UoG: M = 0.439 (SD = 0.22), threshold = 0.89, specificity = 0.709, sensitivity = 0.829).

Overall, there was acceptable agreement between the approaches (Lilly kappa = 0.678; UoG kappa = 0.575, see also Table 2). Although there were instances where PET-anchored measures were negative while distribution-based measures were positive, the inverse was not observed; there were no instances of negative distribution-based pTau-217 with positive PET-anchored pTau-217.

**Table 2.**
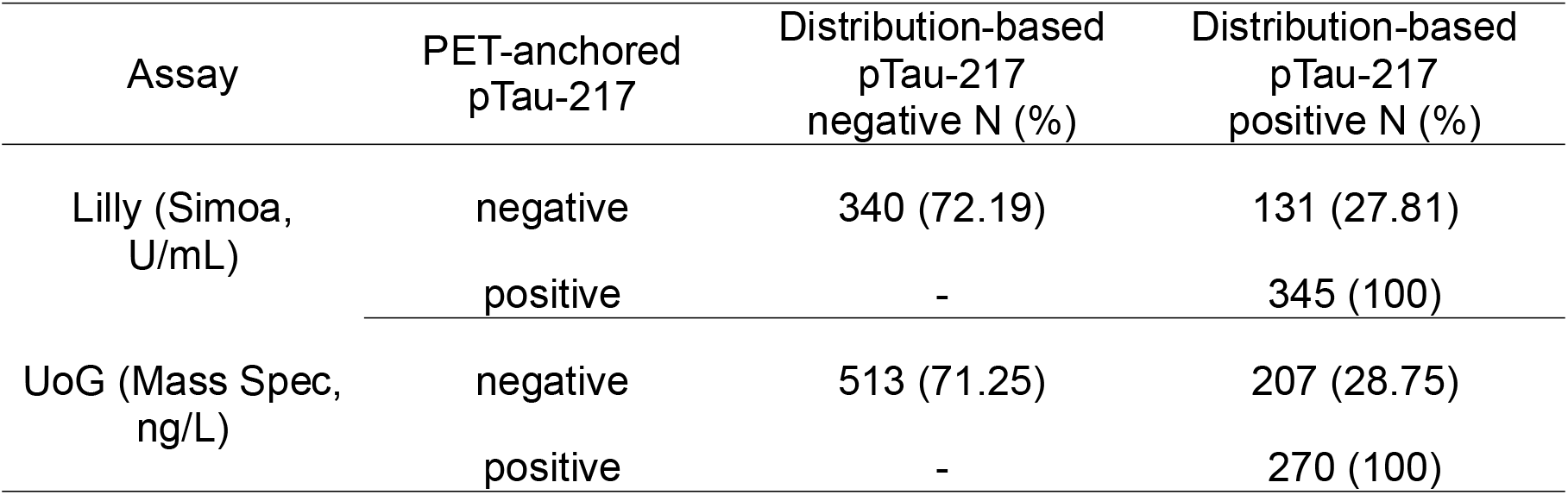
Cross-tabulated PET-anchored and distribution-based pTau-217 positivity.

This disagreement was nearly entirely confined to latent class 2. Both approaches yield the same pattern of latent class 1 being completely pTau-217 negative while class 3 is entirely negative (Figure 3 and Supplementary Material). Class 2 is distinct, in being predominantly pTau-217 negative when PET-anchored and almost exclusively positive when the distribution-based threshold is implemented.

**Figure 3.**
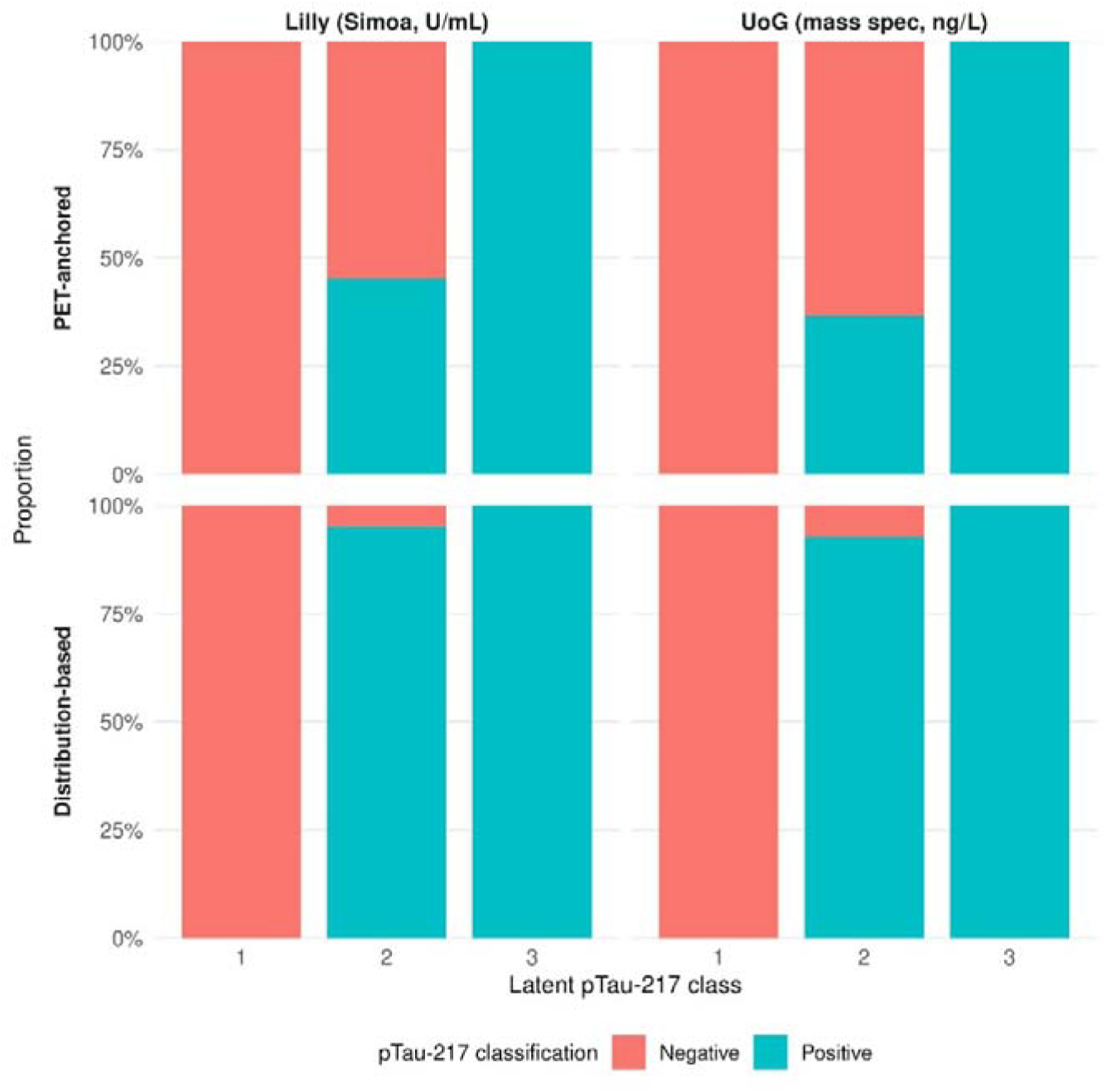
Proportion of latent class by pTau-217 positivity for each assay. *HC* = Healthy Cognition; *MCI* = Mild Cognitive Impairment; *AD* = Alzheimer’s Disease.

## Discussion

We operationalised pTau-217 positivity by anchoring it to AB PET positivity or the observed distribution of concentration. Using an immunoassay and a mass spectrometry assay, we compared the two operationalisation approaches. Both PET-anchored, and distribution-based thresholds produced acceptable overall agreement. However, disagreement was observed in capturing latent biomarker state; classes 1 and 3 were captured similarly, but percentage classified as “positive” differed between approaches. These findings suggest that operational definitions influence the biological interpretation of intermediate pTau-217 concentrations, rather than simply altering the numerical threshold for positivity.

The intermediate latent biomarker class provides insight into the biological heterogeneity underlying plasma pTau-217 concentrations. There was a monotonic increase in both AD diagnosis and AB PET positivity from class 1 to the intermediate class 2, and class 3. The intermediate class itself contains a mix of diagnostic profiles, a different proportion of which was pTau-217 positive depending on benchmarking on its own distribution or AB PET outcome; majority positive when grounded in the biomarker’s own distribution compared to a more equal split when anchored to AB PET. This may represent a state on the biological continuum situated between clearly negative and clearly positive biomarker profiles^4,27,28^, aligned with increasing recognition that pTau-217 reflects a heterogeneous continuum of pathology instead of a binary state ^4,7,29^. Nonetheless, longitudinal or prospective evidence will be required to ascertain whether the intermediate latent reflects disease progression or a stable intermediate state^28,30^.

Misalignment between the thresholding approaches was almost entirely confined to the intermediate latent class, which contextualises the agreement in classifying the other two classes as pTau-217 negative (class 1) or positive (class 3). The PET-anchored threshold is inherently calibrated to pTau-217 profiles that are strongly aligned with brain-derived amyloid pathology evidence, as is currently common with in vivo measures of AD pathology before clinical implementation^7,14^. Contrastingly, the distribution-based threshold has comparatively lower amyloid specificity in favour of sensitivity – thus labelling more of class 2 as positive. Therefore, the two approaches appear to prioritise different biological characteristics, rather than differing only in numerical stringency. This observation contextualises recent work proposing a pTau-217 dual thresholding approach compatible with a biological “grey zone”^28,31^ that accounts for uncertainty. Our findings suggest that such intermediate pTau-217 classifications may indeed reflect a reproducible latent biomarker state.

The discrepancy between operational approaches far outweighed differences between the assays themselves, in alignment with recent evidence of improving analytical harmonisation across pTau-217 platforms, independent of absolute concentration scales^1,7^. Presently observed latent classes were of identical number and with remarkably similar makeup for both the Lilly and UoG assay. In both cases, the intermediate class was the only source of misalignment between operational approaches and the overlap with clinical and AB PET classification was very similar. These findings suggest that operationalisation had greater influence on pTau-217 classification than analytical platform. The reproducibility of this pattern across methodologically distinct assays supports that disagreement between thresholding approaches does not reflect assay-specific behaviour^29^, rather an underlying feature of the pTau-217 biomarker continuum.

The practical significance of these findings lies in the interpretation of biomarker positivity, which is not a fixed property independent of its operational definition. Ultimately, biomarker positivity reflects the biological reference against which it is calibrated. PET-anchored thresholds prioritise alignment with cerebral amyloid plaque accumulation^5^, consistent with recommendation for emerging biomarkers to be interpreted relative to established biological references before clinical implementation^14^. Distribution-based thresholds instead identify a broader region of the plasma pTau-217 continuum, potentially capturing cases that may not yet have reached PET-defined amyloid positivity. This interpretation is consistent with growing evidence that plasma pTau-217 changes can be heterogeneous and occur before severe amyloid plaque accumulation or overt clinical progression. Indeed, contemporary models increasingly recognise AD as a dynamic pathology continuum, rather than a dichotomous state, further reinforcing the importance of selecting the most appropriate thresholding approach for the intended purpose.

Important caveats to these results relate to the nature of the cohort dataset, as cross-sectional without longitudinal follow-up that could explore temporal and disease progression effects. Moreover, brain-derived evidence of tau accumulation is not available to deepen understanding of each threshold’s meaning. Future longitudinal studies should determine the alignment of thresholding approaches in reference to clinical classification, brain-derived pathogenic staging, and overall disease staging. Presently reported thresholds were derived and evaluated within the same sample, which future research could expand on using additional datasets for external validation.

In summary, PET-anchored and distribution-based thresholds for pTau-217 positivity produced comparable classifications at the extremes of the continuum. The approaches diverged in classifying an intermediate biomarker state. These patterns were consistent across two independent and distinct analytical platforms. These findings suggest that operational definitions of pTau-217 positivity shape the state on the biological continuum reflected in the resulting classification. Selection of thresholding strategies should therefore be considered a methodological, rather than technical, decision tailored to the intended application and target biological state.

## Statements

### Ethics

This study was part of the University of Glasgow-led Data Challenge, with ethical approval by the University of Glasgow Medical, Veterinary, and Life Sciences College ethics committee (number 200230198).

### Data availability

The Bio-Hermes-001 dataset (Clinical Trials ID: NCT04733989) is currently hosted on Alzheimer’s Disease Data Initiative (ADDI, report doi: 10.1002/alz.70278)), where it can be accessed upon request (https://discover.alzheimersdata.org/catalogue/datasets/8395cc96-f249-40db-beaa-4668feb3cc5e).

### Generative AI statement

In compliance with Wiley guidance for generative AI use, ChatGPT (OpenAI, v 5.5) was used to support manuscript structuring and editing to improve clarity and readability. During formal analysis, it was used to streamline code written by the research team. The authors take full responsibility for all scientific content and the final manuscript.

### Author contributions

KM: conceptualisation; data curation; formal analysis; investigation; methodology; project administration; resources; software; visualisation; writing – original draft; writing – review & editing.

AD: methodology; writing – review & editing

JB: methodology; writing – review & editing

LH: resources; writing – review & editing

SK: methodology; writing – review & editing

TQ: supervision; writing – review & editing

