## Supplementary Material for "Latent biomarker states underlying disagreement between PET-anchored and distribution-based plasma pTau-217 positivity thresholds"

-

Supplementary Material

**Corresponding author:**

Kalliopi Mavromati

**Results**

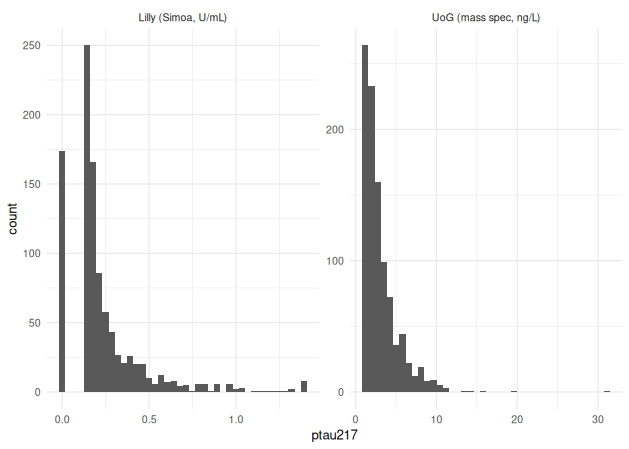
**Figure S1.** Distribution of non-transformed pTau-217 by assay.

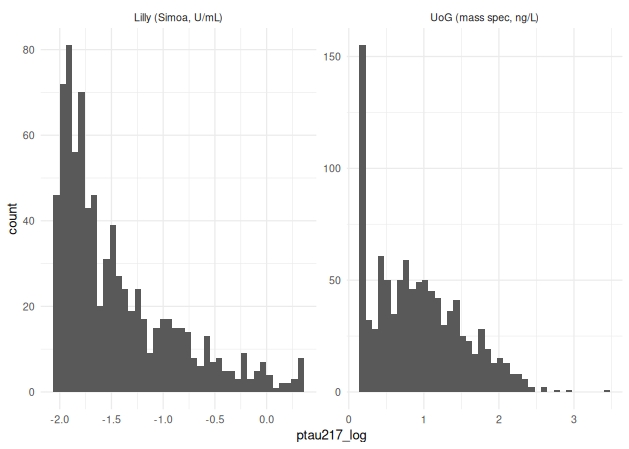
**Figure S2.** Distribution of log-transformed pTau-217 by assay.

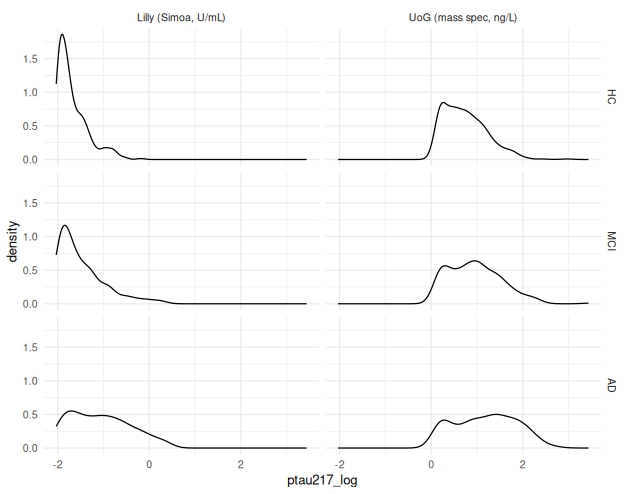
**Figure S3.** Distribution of log-transformed pTau-217 by assay and clinical diagnosis. *HC* = Healthy Cognition; *MCI* = Mild Cognitive Impairment; *AD* = Alzheimer’s Disease.

**Figure S4.** Distribution of log-transformed pTau-217 by sex. *F* = Female; *M* = Male.
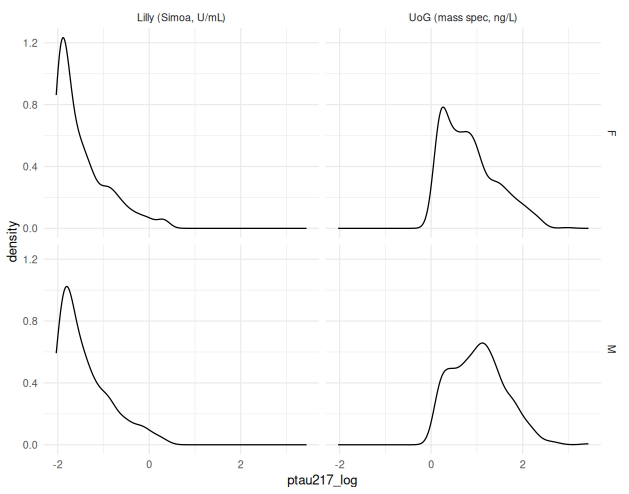

| **Table S1.** Count and proportion of latent class by clinical diagnosis and AB PET positivity for each assay. | | | | | | | |
| --- | --- | --- | --- | --- | --- | --- | --- |
| Assay | Latent Class | HC N (%) | MCI N (%) | AD N (%) | AB PET negative N (%) | AB PET positive N (%) | AB PET missing N (%) |
| Lilly (Simoa, U/mL) | 1 | 187 (57.19) | 98 (29.97) | 42 (12.84) | 281 (85.93) | 28 (8.56) | 18 (5.50) |
|  | 2 | 106 (40.30) | 96 (36.50) | 61 (23.19) | 137 (52.09) | 115 (43.73) | 11 (4.18) |
|  | 3 | 35 (15.49) | 58 (25.66) | 133 (58.85) | 23 (10.18) | 190 (84.07) | 13 (5.75) |
| UoG (Mass Spec, ng/L) | 1 | 259 (53.29) | 140 (28.81) | 87 (17.90) | 404 (83.13) | 51 (10.49) | 31 (6.38) |
|  | 2 | 137 (37.13) | 129 (34.96) | 103 (27.91) | 173 (46.88) | 181 (49.05) | 15 (4.06) |
|  | 3 | 20 (14.81) | 39 (28.89) | 76 (56.30) | 20 (14.81) | 107 (79.26) | 8 (5.92) |

*Note.* *HC* = Healthy Cognition; *MCI* = Mild Cognitive Impairment; *AD* = Alzheimer’s Disease; *AB PET* = amyloid beta PET.

| **Table S2.** Count and proportion of latent class pTau-217 positive for each thresholding approach and assay. | | | | | |
| --- | --- | --- | --- | --- | --- |
| Assay | Latent class | PET-anchored pTau-217 negative N (%) | PET-anchored pTau-217 positive N (%) | reference approach pTau-217 negative N (%) | reference approach pTau-217 positive N (%) |
| Lilly (Simoa, U/mL) | 1 | 327 (100) | - | 327 (100) | - |
|  | 2 | 144 (54.75) | 119 (45.25) | 13 (4.94) | 250 (95.06) |
|  | 3 | - | 226 (100) | - | 226 (100) |
| UoG (Mass Spec, ng/L) | 1 | 486 (100) | - | 486 (100) | - |
|  | 2 | 234 (63.41) | 135 (36.58) | 27 (7.32) | 342 (92.68) |
|  | 3 | - | 135 (100) | - | 135 (100) |

*Note.* – means there was no prevalence of specified condition.
